# Impact of vaccination and undetected cases on the COVID-19 pandemic dynamics in Qatar in 2021

**DOI:** 10.1101/2021.05.27.21257929

**Authors:** Igor Nesteruk

**Affiliations:** Institute of Hydromechanics, National Academy of Sciences of Ukraine National Technical University of Ukraine “Igor Sikorsky Kyiv Polytechnic Institute”

**Keywords:** COVID-19 pandemic, vaccination efficiency, epidemic dynamics in Qatar, mathematical modeling of infection diseases, SIR model, parameter identification, statistical methods

## Abstract

The third COVID-19 pandemic wave in Qatar was simulated with the use of the generalized SIR-model and the accumulated number of cases reported by Johns Hopkins University for the period: April 25 - May 8, 2021. Comparison with the SIR-curves calculated before for the second wave showed that the effect of mass vaccination is not evident during 4 months after its onset in December 2020. Additional simulations have demonstrated that many COVID-19 cases are not detected. The real accumulated number of cases can exceed the laboratory-confirmed one more than 5 times. This fact drastically increases the probability of meeting an infectious person and the epidemic duration.

## Introduction

The COVID-19 pandemic dynamics in Qatar was simulated with the use of SEIR-model (susceptible-exposed-infected-removed) [1] and SEIRD-model (susceptible-exposed-infected-removed-dead) [2]. Some recent SIR-simulations [3] were based on the dataset about the number of cases in December 2020, when the mass vaccination started in this country. In this study we will use the information about the accumulated number of cases from COVID-19 Data Repository by the Center for Systems Science and Engineering (CSSE) at Johns Hopkins University (JHU), [4]. We will analyze the recent epidemic dynamics in Qatar and make some predictions taking into account the incompleteness of the statistical information with the use of method proposed in [5].

### Data

We will use two data sets regarding the accumulated numbers of confirmed COVID-19 cases *V*_*j*_, number of vaccinated people *S*_*j*_ and number of vaccinations *Q*_*j*_ in Qatar from JHU, [4]. These values and corresponding moments of time *t*_*j*_ (measured in days) are shown in Table 1. For SIR-simulations we have used the values of *V*_*j*_ and *t*_*j*_ corresponding to the time period *T*_*c3*_ : April 25 – May 8, 2021 during the third epidemic wave in Qatar. Other values presented in Table 1 and datasets available in [3] were used only for comparisons and verifications of the calculations.

**Table 1.**
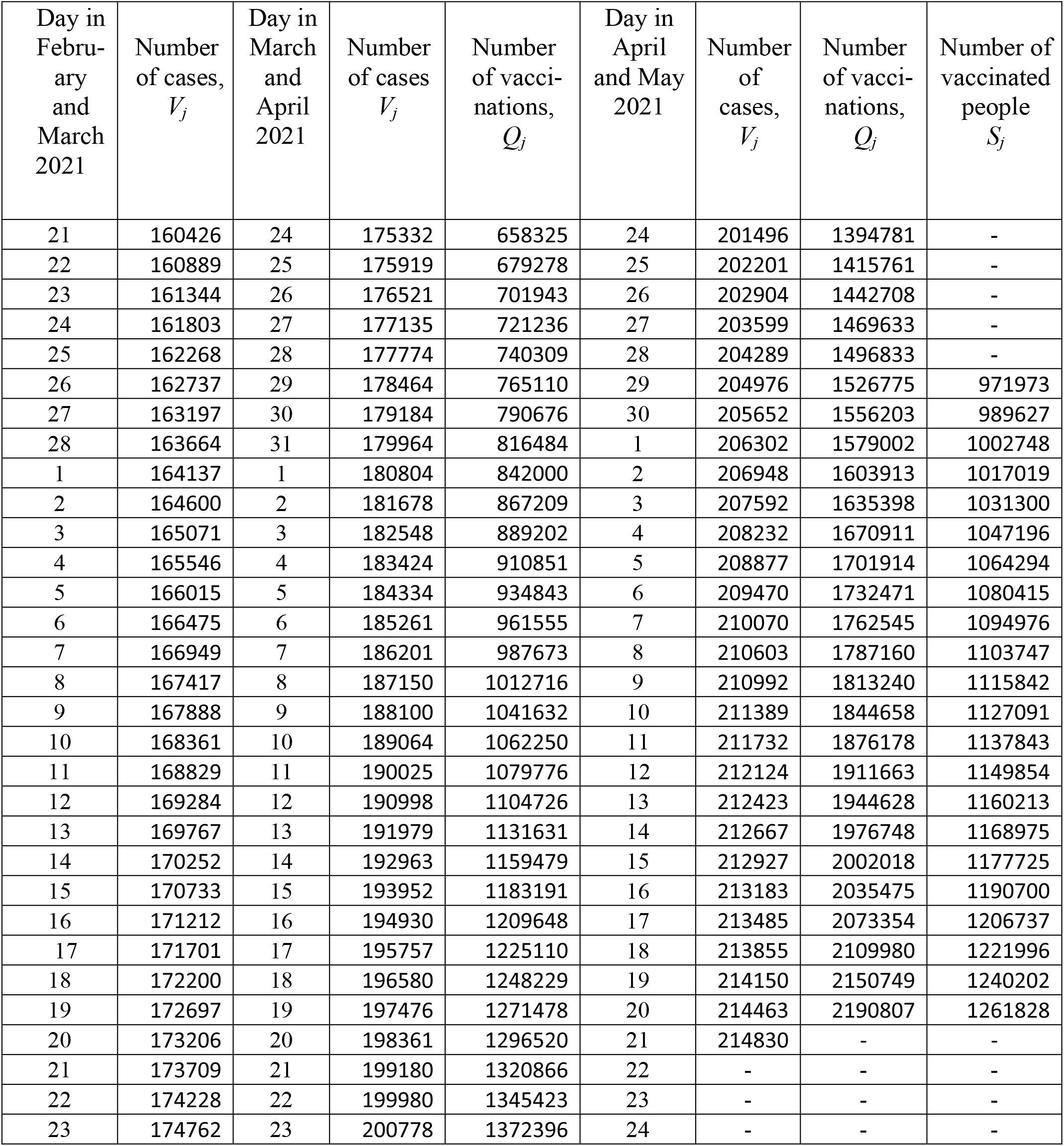
Cumulative numbers for Qatar from [4]. The laboratory-confirmed Covid-19 cases *V*_*j*_, number of vaccinations *Q*_*j*_, the number of vaccinated people *S*_*j*_, and corresponding moments of time.

| Day in February and March 2021 | Number of cases, $V_j$ | Day in March and April 2021 | Number of cases $V_j$ | Number of vaccinations, $Q_j$ | Day in April and May 2021 | Number of cases, $V_j$ | Number of vaccinations, $Q_j$ | Number of vaccinated people $S_j$ |
| --- | --- | --- | --- | --- | --- | --- | --- | --- |
| 21 | 160426 | 24 | 175332 | 658325 | 24 | 201496 | 1394781 | - |
| 22 | 160889 | 25 | 175919 | 679278 | 25 | 202201 | 1415761 | - |
| 23 | 161344 | 26 | 176521 | 701943 | 26 | 202904 | 1442708 | - |
| 24 | 161803 | 27 | 177135 | 721236 | 27 | 203599 | 1469633 | - |
| 25 | 162268 | 28 | 177774 | 740309 | 28 | 204289 | 1496833 | - |
| 26 | 162737 | 29 | 178464 | 765110 | 29 | 204976 | 1526775 | 971973 |
| 27 | 163197 | 30 | 179184 | 790676 | 30 | 205652 | 1556203 | 989627 |
| 28 | 163664 | 31 | 179964 | 816484 | 1 | 206302 | 1579002 | 1002748 |
| 1 | 164137 | 1 | 180804 | 842000 | 2 | 206948 | 1603913 | 1017019 |
| 2 | 164600 | 2 | 181678 | 867209 | 3 | 207592 | 1635398 | 1031300 |
| 3 | 165071 | 3 | 182548 | 889202 | 4 | 208232 | 1670911 | 1047196 |
| 4 | 165546 | 4 | 183424 | 910851 | 5 | 208877 | 1701914 | 1064294 |
| 5 | 166015 | 5 | 184334 | 934843 | 6 | 209470 | 1732471 | 1080415 |
| 6 | 166475 | 6 | 185261 | 961555 | 7 | 210070 | 1762545 | 1094976 |
| 7 | 166949 | 7 | 186201 | 987673 | 8 | 210603 | 1787160 | 1103747 |
| 8 | 167417 | 8 | 187150 | 1012716 | 9 | 210992 | 1813240 | 1115842 |
| 9 | 167888 | 9 | 188100 | 1041632 | 10 | 211389 | 1844658 | 1127091 |
| 10 | 168361 | 10 | 189064 | 1062250 | 11 | 211732 | 1876178 | 1137843 |
| 11 | 168829 | 11 | 190025 | 1079776 | 12 | 212124 | 1911663 | 1149854 |
| 12 | 169284 | 12 | 190998 | 1104726 | 13 | 212423 | 1944628 | 1160213 |
| 13 | 169767 | 13 | 191979 | 1131631 | 14 | 212667 | 1976748 | 1168975 |
| 14 | 170252 | 14 | 192963 | 1159479 | 15 | 212927 | 2002018 | 1177725 |
| 15 | 170733 | 15 | 193952 | 1183191 | 16 | 213183 | 2035475 | 1190700 |
| 16 | 171212 | 16 | 194930 | 1209648 | 17 | 213485 | 2073354 | 1206737 |
| 17 | 171701 | 17 | 195757 | 1225110 | 18 | 213855 | 2109980 | 1221996 |
| 18 | 172200 | 18 | 196580 | 1248229 | 19 | 214150 | 2150749 | 1240202 |
| 19 | 172697 | 19 | 197476 | 1271478 | 20 | 214463 | 2190807 | 1261828 |
| 20 | 173206 | 20 | 198361 | 1296520 | 21 | 214830 | - | - |
| 21 | 173709 | 21 | 199180 | 1320866 | 22 | - | - | - |
| 22 | 174228 | 22 | 199980 | 1345423 | 23 | - | - | - |
| 23 | 174762 | 23 | 200778 | 1372396 | 24 | - | - | - |

### Generalized SIR model and parameter identification procedure

The classical SIR model for an infectious disease [6-8] was generalized in [5, 9] to simulate different epidemic waves. We suppose that the SIR model parameters are constant for every epidemic wave, i.e. for the time periods: 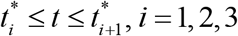 Then for every wave we can use the equations, similar to [6-8]:

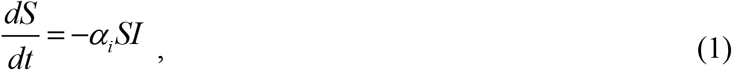

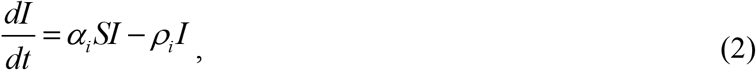

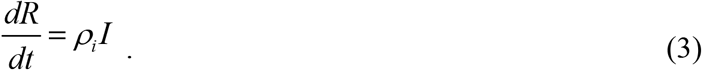

Here *S* is the number of susceptible persons (who are sensitive to the pathogen and **not protected**); *I* is the number of infectious persons (who are sick and **spread the infection)**; and *R* is the number of removed persons (who **no longer spread the infection)**. It must be noted that *I(t)* is not the number of active cases. People can be ill (among active cases), but isolated. In means, that they don’t spread the infection anymore. There are many people spreading the infection but not tested and registered as active cases. The use of number of active cases as *I(t)* in some papers a principal mistake which may lead to incorrect results. Parameters *α*_*i*_ and *ρ*_*i*_ are supposed to be constant for every epidemic wave.

To determine the initial conditions for the set of equations (1)–(3), let us suppose that at the beginning of every epidemic wave 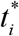 :

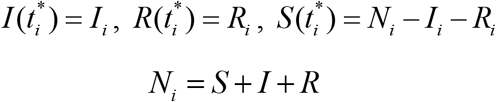

In [5, 9] the set of differential equations (1)-(3) was solved by introducing the function

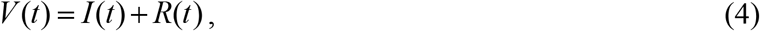

corresponding to the number of victims or the cumulative number of cases. The corresponding analytical formulas for this exact solution; the saturation levels *S*_*i*∞_ ; *V*_*i*∞_ = *N*_*i*_ − *S*_*i*∞_ (corresponding the infinite time moment) and the final day of the *i-th* epidemic wave (corresponding the moment of time when the number of persons spreading the infection will be less than 1) can be found in [5, 9].

For many epidemics (including the COVID-19 pandemic) we cannot observe dependencies *S*(*t*), *I* (*t*) and *R*(*t*) but observations of the accumulated number of cases *V*_*j*_ corresponding to the moments of time *t*_*j*_ provide information for direct assessments of the dependence *V* (*t*). In the case of a new epidemic, the values of its parameters are unknown and must be identified with the use of limited data sets. For the second and next epidemic waves (*i* > 1), the moments of time 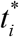 corresponding totheir beginning are known. Therefore the exact solution [5, 9] depend only on five parameters - *N*_*i*_, *I*_*i*_, *R*_*i*_, *v*_*i*_, *α*_*i*_, when the registered number of victims *V*_*j*_ is a random realization of its theoretical dependence (4).

The real number of COVID-19 cases is much higher than the number of laboratory-confirmed patients [5, 10-15], since many patients have no symptoms or make no tests. If we assume, that data set *V*_*j*_ is incomplete and there is a constant coefficient *β*_*i*_ ≥ 1, relating the registered and real number of cases during the *i-th* epidemic wave:

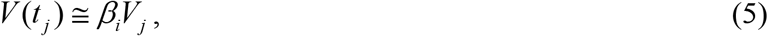

then the number of unknown parameters increases by one. The values *V*_*j*_, corresponding to the moments of time *t*_*j*_, the exact solution [5, 9] and relationship (5) can be used to find the optimal values of the parameters *α*_*i*_, *β*_*i*_, *N*_*i*_, *v*_*i*_, *I*_*i*_, *R*_*i*_ providing the maximum value of the correlation coefficient *r*_*i*_ (see details in [5, 16]).

## Results

First we have calculated the optimal vales of parameters for the third wave of the COVID-19 pandemic in Qatar assuming that the dataset *V*_*j*_ and *t*_*j*_ reflects the real number of cases (i.e., we supposed that *β*_3_ = 1). The results are shown in Table 2 (third column). The last column of the Table 2 represents the characteristics of the second wave calculated in [3] for *β*_2_ = 1. It can be seen that the optimal values of parameters *N*_*i*_, *v*_*i*_, *α*_*i*_ and the final sizes *V*_*i*∞_ are rather different for the second an third pandemic waves in Qatar (see last two columns in Table 2), but the estimations of average time of spreading the infection 1/ *ρ*_*i*_ ≈ 3.9 days and the epidemic duration are very close. The corresponding SIR curves are shown in Fig. 1 by black (wave 2, *β*_2_ =1) and red (wave 3, *β*_3_ =1) colors. Solid lines represent the number of victims *V(t)=R(t)+I(t)*, dashed lines – the number of infectious persons *I(t)* and dotted lines - the derivatives

**Table 2.** Calculated optimal values of SIR parameters and other characteristics of the second and third pandemic waves in Qatar.

| Characteristics | Invisible wave 3,<br>$i=3$,<br>$\beta_3 = 5.308$ | Visible wave 3,<br>$i=3$,<br>$\beta_3 = 1$ | Visible wave 2, [3]<br>$i=2$,<br>$\beta_2 = 1$ |
| --- | --- | --- | --- |
| Period taken for calculations, $T_{ci}$ | April 25 – May 8, 2021 | April 25 – May 8, 2021 | December 11-24, 2020 |
| $I_i$ | 15,895.4703455678 | 2,993.09405674162 | 581.6852057918 |
| $R_i$ | 1,057,266.11194015 | 199,185.048800401 | 140,084.171937066 |
| $N_i$ | 3,102,717.088 | 571,480 | 343,800 |
| $\nu_i$ | 2,137,169.15821761 | 388,684.616416235 | 203,678.101450066 |
| $\alpha_i$ | 1.18748428868345e-07 | 6.52942943554382e-07 | 1.26938469796243e-06 |
| $\rho_i$ | 0.253785479764225 | 0.253788877557122 | 0.258545865290753 |
| $1/\rho_i$ | 3.94033575494167 | 3.94028300068005 | 3.86778569781199 |
| $r_i$ | 0.999979810302661 | 0.99997979869081 | 0.999943364258809 |
| $S_{i\infty}$ | 1,866,781 | 338,877 | 188,661 |
| $V_{i\infty}$ | 1,235,936 | 232,603 | 155,139 |
| Final day of the epidemic wave | March 14, 2022 | January 18, 2022 | January 16, 2022 |

**Fig. 1.**
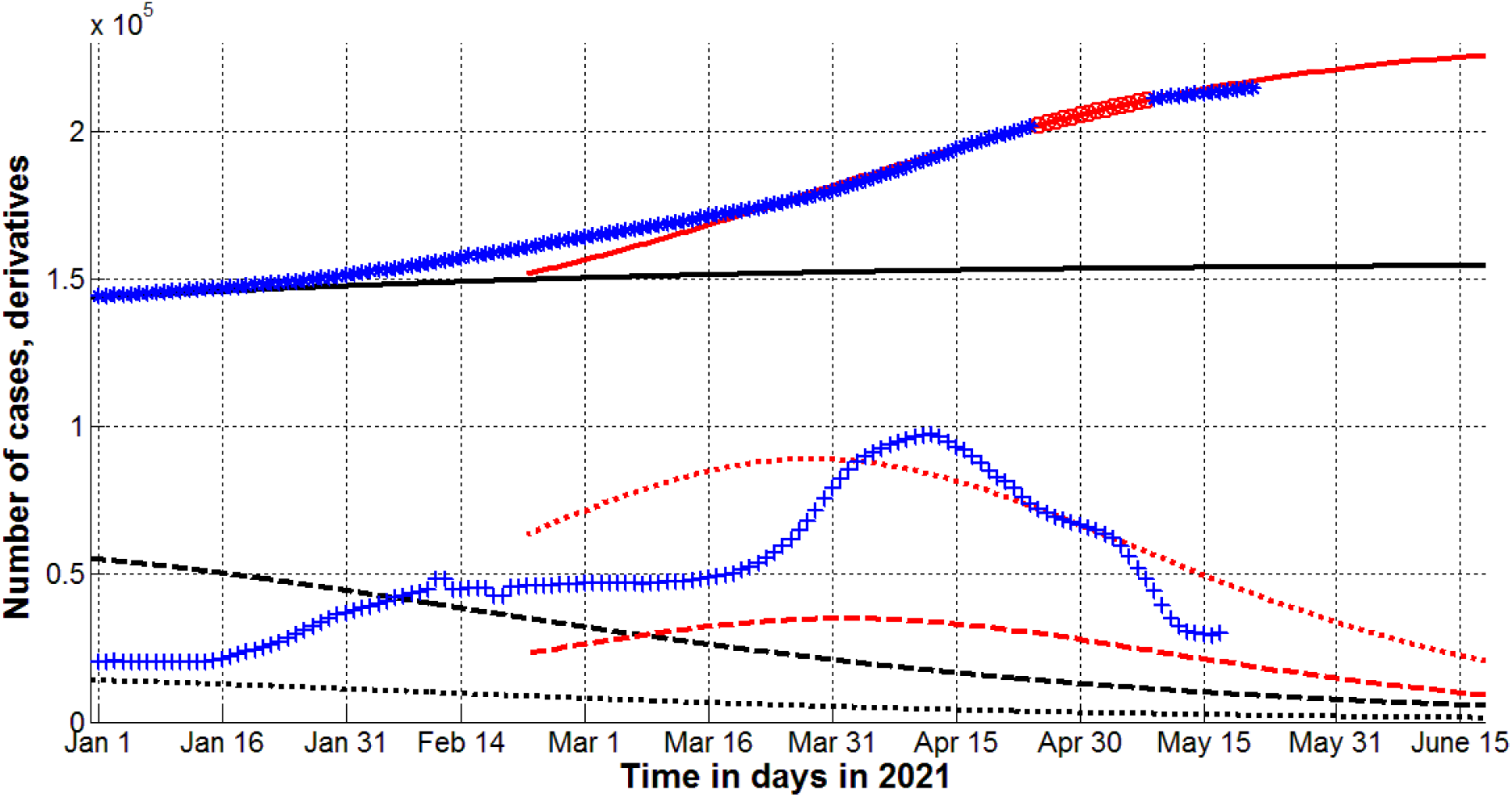
Visible second (black, [3]) and third (red) pandemic waves in Qatar. SIR simulations (lines) calculated at *β*_*i*_ =1. Numbers of victims *V(t)=I(t)+R(t)* – solid lines; numbers of infected and spreading *I(t)* multiplied by 10 – dashed; derivatives *dV/dt* (eq. (6)) multiplied by 100 – dotted. Markers show accumulated numbers of laboratory-confirmed cases *V*_*j*_ (from Table 1 and [3]) and derivatives. Red “Circles” correspond to the accumulated numbers of cases taken for calculations of the third wave (during period of time *T*_*c3*_); blue “stars” – number of cases beyond *T*_*c3*_. “Crosses” show the first derivatives (eq. (8)) multiplied by 100.

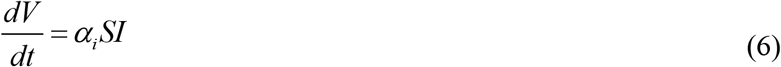

Equation (6) follows from (2)-(4) and yields a theoretical estimation of the average daily number of new cases which can be calculated by numerical differentiation of the smoothed accumulated number of cases

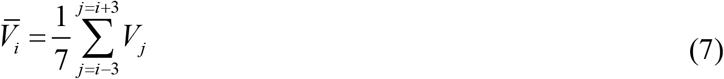

as follows:

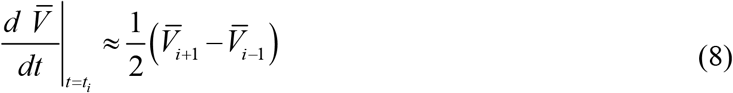

Values of the derivative (8) are shown in Fig. 1 by blue crosses and illustrate that the experimental and theoretical estimation for the second epidemic wave (eq. (6), dotted black line) started to deviate after January 16, 2021. At the end of March 2021 there was a sharp increase of the averaged daily number of new cases. In April 2021 derivative (8) was close to the theoretical estimation (6) for the third wave (compare crosses and the red dotted line in Fig. 1).

Blue “stars” in Fig. 1 show the laboratory-confirmed cases *V*_*j*_ (from Table 1 and corresponding Table in [3]) used only to check the results of calculation (in comparison with red “circles” corresponding to values taken for SIR simulations of the third epidemic wave in Qatar). It can be seen that the registered number of cases deviates from the theoretical curve for the second wave (compare the black line and “stars” in Fig. 1). The *V(t)=I(t)+R(t)* curve for the third wave (calculated with the use of fresher dataset) is in good agreement with the accumulated number of cases confirmed after mid-March 2021 (compare the red line and “stars” in Fig. 1).

Assuming that the dataset *V*_*j*_ does not reflect the real number of cases, we have calculated the optimal value of the visibility coefficient *β*_3_ = 5.308 in relationship (5) and other optimal vales of parameters for the third wave of the COVID-19 pandemic in Qatar. The results are shown in Table 2 (second column). It can be seen that the correlation coefficient for the invisible wave 3 is higher than for its visible part. There is a huge difference in the optimal values of parameters *N*_*i*_, *v*_*i*_, *α*_*I*_ and the final sizes *V*_*i*∞_ ; large difference in the epidemic duration, but the estimations of average time of spreading the infection 1/ *ρ*_*i*_ ≈ 3.9 days are very close (compare second and third column in Table 2).

Red lines in Fig.2 represent the SIR curves for the invisible dynamics of the third epidemic wave in Qatar. Solid line shows the number of victims *V(t)=R(t)+I(t)*, dashed line – the number of infectious persons *I(t)* multiplied by 10 and dotted line - the derivative (6) multiplied by 100. The accumulated numbers the laboratory-confirmed cases *V*_*j*_ (shown by blue “stars” and red “circles”) are much lover than the theoretical estimation of real number of cases *V(t)=R(t)+I(t)* (shown by the solid red line).

To check the reliability of the method and the results of calculations the accumulated number of laboratory-confirmed cases *V*_*j*_ (listed in Table 1) was smoothed with use of formula (7) and corresponding values 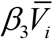 are shown by the blue line. It can be seen that theoretical estimation (the red line) is very close the recorded number of cases multiplied by the optimal value of the visibility coefficient *β*_3_ = 5.308. The values of the derivative of the smoothed accumulated number of laboratory-confirmed cases (calculated with the use of equations (7) and (8)) have been multiplied by the optimal value of the visibility coefficient *β*_3_ = 5.308 and shown in Fig. 2 by blue crosses which are close to the theoretical dotted line in April 2021.

**Fig. 2.**
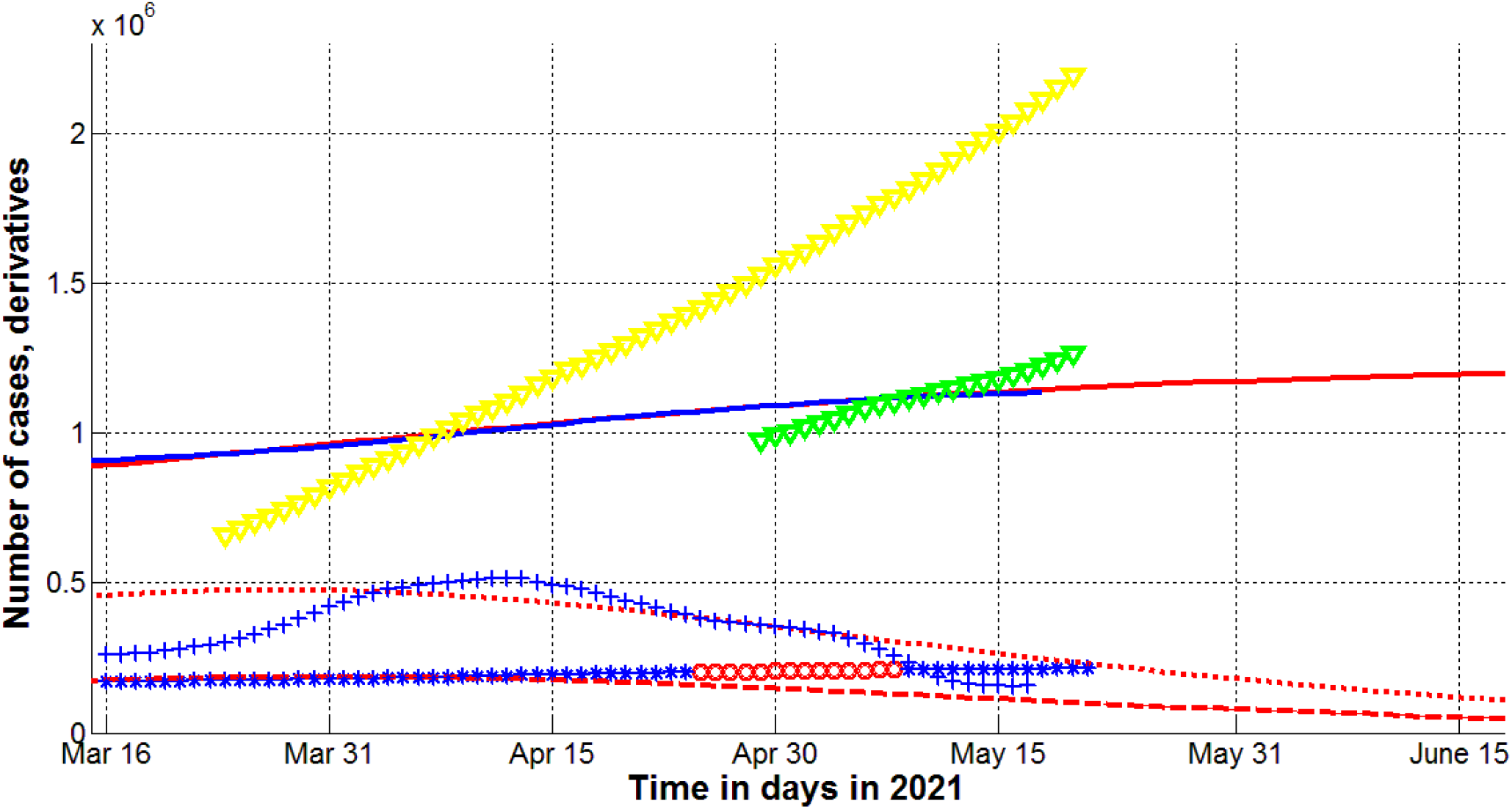
Real sizes of the third pandemic wave in Qatar for the calculated optimal value of the visibility coefficient *β*_3_ = 5.308. SIR simulations (red lines), numbers of vaccinations (yellow) and vaccinated persons (green). Numbers of victims *V(t)=I(t)+R(t)* – solid lines; numbers of infected and spreading *I(t)* multiplied by 10 – dashed; derivatives *dV/dt* (eq. (6)) multiplied by 100 – dotted. Markers show accumulated numbers of laboratory-confirmed cases *V*_*j*_ (from Table 1) and derivatives. Red “Circles” correspond to the accumulated numbers of cases taken for calculations of the third wave (during period of time *T*_*c3*_); blue “stars” – number of cases beyond *T*_*c3*_. “Crosses” show the first derivatives (eq. (8)) multiplied by 100*β*_3_. The blue line represents smoothed accumulated number of laboratory-confirmed cases (7) multiplied by *β*_3_.

## Discussion

The mass vaccination started in Qatar in late December 2020. The corresponding numbers of vaccinated people *S*_*j*_ and vaccinations *Q*_*j*_ are shown in Fig. 2 by “green” and “yellow” markers, respectively. Despite the relatively rapid rate of vaccination (in May 2021, the number of fully vaccinated people *Q*_*j*_ - *S*_*j*_ approached half of the population), Qatar experienced the new wave of pandemics with a sharp increase in the number of patients in March-April 2021 (see blue “crosses” in Fig. 1). According to the forecast of the previous second wave (made in [3] using data before vaccination), the number of cases of should stabilize rapidly in 2021 (see the black solid line in Fig. 1). We will probably see the effect of vaccination only in June 2021. At least the latest data on the daily number of new cases have already become less than the theoretical estimates for the third wave (compare “blue” crosses and red dotted lines in Figs. 1 and 2).

The calculated coefficient of epidemic visibility *β*_3_ = 5.308 for Qatar correlates with the values *β*_9_ =4.1024 and *β*_10_ =3.7 obtained in [5] for the ninth and tenth epidemic waves in Ukraine and the results of random testing in two kindergartens and two schools in the Ukrainian city of Chmelnytskii [15] which revealed the value 3.9. The total testing in Slovakia (65.5% of population was tested on October 31-November 1, 2020) revealed a number of previously undetected cases, equal to about 1% of the population [13]. On November 7 next 24% of the population was tested and found 0.63% of those infected [14]. According to the WHO report at the end of October, the number of detected cases in Slovakia was also approximately 1% of population [17].

Ignoring information about the large number of unreported cases can lead to incorrect recommendations for quarantine restrictions and overly optimistic forecasts of the COVID-19 pandemic duration. For example, the information about the real dependence *I(t)* is important to estimate the probability of meeting an infected person with the use of simple formula, [18]:

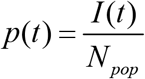

Where *N*_*pop*_ is the volume of population. As of the end of May, 2021 the theoretical estimation yields the value *I* ≈ of 10,000 (see the dashed line in Fig. 2). Then the probability *p* can be estimated as 0.0034 for Qatar. If only officially registered cases are taken into account, the corresponding probability will be approximately 5.3 times lower.

If current trends continue, new cases in Qatar will cease to be registered in January 2022 (see the third column in Table 2). But the invisible cases will continue for another two months (see the second column in Table 2). During this long period, new strains of the coronavirus may emerge and cause a new epidemic wave, which will be unexpected, as the visible part of the epidemic seemed to have been overcome.

## Conclusions

The application of generalized SIR model to the new COVID-19 pandemic wave in Qatar demonstrated once more its effectiveness in predictions of epidemic dynamics and estimations of the vaccination efficiency. The parameter identification procedure allowed calculating the coefficients of data incompleteness (approximately 5.3 in the end of April 2021). New simulations with the use of fresher datasets are necessary to update the estimation of the vaccination efficiency in Qatar. Probably, real sizes of the pandemic in other countries are also much large than the number of registered cases. Thus reassessments of the COVID-19 pandemic dynamics are necessary, to avoid new unexpected waves.

## Data Availability

All data is in text

## Acknowledgements

The author is grateful to Oleksii Rodionov for his help in collecting and processing data.

